# Self-reported sleep problems are associated with impaired daily-life gait quality and increased fall risk in older people

**DOI:** 10.64898/2026.03.30.26349800

**Authors:** Kimberley S. van Schooten, Andrew Vakulin, Rajani Khanal, Kelly Sansom, James Bletsas, Kim Delbaere

**Affiliations:** Neuroscience Research Australia, Randwick, New South Wales, Australia; School of Population Health, University of New South Wales, Kensington, New South Wales, Australia; Flinders Health and Medical Research Institute (FHMRI), Sleep Health, Flinders University, Adelaide, South Australia, Australia; Centre for Healthy Ageing, Health Futures Institute, Murdoch University, Murdoch, Western Australia, Australia; School of Biomedical Science, University of New South Wales, Kensington, New South Wales, Australia; School of Health Sciences, University of New South Wales, Kensington, New South Wales, Australia

**Keywords:** Motor control, daily-life mobility, accidental falls, sleep disorders

## Abstract

**Background:** Sleep problems are common in older people and have been associated with increased fall risk, but the mechanisms underlying this relationship remain unclear. Gait quality reflects balance control and neurological function and may provide insight into pathways linking sleep health and falls.

**Methods:** Data from 758 community-dwelling older people (≥65 years; mean age 75.8 years, 69.3% women) were analysed. Sleep problems were assessed at baseline using a self-reported item (Patient Health Questionnaire-9, question 3). Daily-life gait quality and habitual walking speed were derived from one week of wearable sensor monitoring. Falls and injurious falls were prospectively recorded over 12 months. Associations between sleep problems, gait quality, and fall incidence were examined using regression models adjusted for demographic, pain and cognitive factors, and use of sleeping medication.

**Results:** Sleep problems were reported by 43.9% of participants. Sleep problems were not associated with habitual walking speed, but were associated with lower gait quality in daily life (adjusted β = −0.15, 95% CI −0.27 to −0.03). Participants reporting sleep problems had higher incidence rates of total falls (adjusted IRR = 1.42, 95% CI 1.07 to 1.90) and injurious falls (adjusted IRR = 1.50, 95% CI 1.07 to 2.10).

**Conclusions:** Self-reported sleep problems were associated with impaired real-world gait quality and substantially higher rates of falls and injurious falls in older people. These findings suggest that sleep problems may increase fall risk by altering balance control rather than by reducing walking speed. Sleep should be considered when managing fall risk, and fall risk should be considered in older people with sleep complaints.

**KEY POINTS BOX:** *Key points:*

- Sleep problems in older people were not associated with habitual walking speed, but were associated with lower gait quality in daily life.
- People reporting sleep problems had 42% higher rates of falls, and 50% higher rates of injurious falls.

*Why does this paper matter?:* This paper highlights that sleep problems are an important and under-recognised marker of fall risk among older people. It advocates for the need to consider sleep in fall risk management and fall risk in those presenting with sleep complaints.

## INTRODUCTION

Falls are a leading cause of injury, disability and loss of independence in older people and present a major public health challenge worldwide ^1^. Every year, about one in three community-living older people experience at least one fall, which have substantial consequences for physical function, quality of life and care utilisation ^2^. Despite extensive research, fall prevention remains challenging, partly because falls arise from the interaction of multiple physiological, cognitive, and behavioural factors that change with ageing and disease ^3^. Improving fall risk prediction and identifying modifiable fall risk factors, therefore, remain key priorities ^4^.

Sleep problems are highly prevalent in older people and increase with age ^5^. Common problems include difficulty initiating or maintaining sleep, early morning awakening, non-restorative sleep, and altered sleep timing ^6^. These changes reflect a combination of age-related neurobiological alterations, medical comorbidities, medication use, and lifestyle factors ^6^. Poor sleep has been linked to a wide range of adverse health outcomes, including cardiovascular disease, metabolic dysfunction, mood disorders, and cognitive decline ^6^. More recently, poor sleep has also been recognised as a potential contributor to mobility impairment and falls, although this relationship remains poorly understood ^7^.

Several pathways may link disturbed sleep to increased fall risk in older people. At the cognitive level, insufficient or poor-quality sleep is associated with reduced attention, slower processing speed, impaired executive function, and diminished dual-task capacity ^8^. These cognitive domains are critical for safe mobility in daily life, where walking rarely occurs in isolation but instead requires continuous adaptation to environmental demands, obstacles, and competing tasks ^9^. Sleep may also influence falls through effects on balance and motor control. Experimental studies have shown that sleep restriction and sleep fragmentation can impair postural stability, increase sway, and alter neuromuscular control ^10,11^. In addition to these direct effects, sleep problems frequently co-occur with other fall-related risk factors, including chronic pain, depression, anxiety, and neurodegenerative disease ^12^. Medications for some of these conditions can also affect balance and alertness ^13^, complicating the interpretation of observed associations. Finally, fatigue and daytime sleepiness may reduce physical activity or alter movement strategies ^14^, potentially influencing exposure to fall risk in everyday life. Disentangling the independent contribution of sleep from these correlated factors is therefore challenging but necessary to clarify its role in fall aetiology.

Most epidemiological studies examining sleep and falls rely on retrospective self-reported falls ^7^, and those between sleep and balance rely on laboratory- or clinic-based mobility assessments, limiting insight into real-world mechanisms and increasing susceptibility to recall bias. Advances in wearable sensor technology have enabled objective assessment of real-world gait characteristics over extended periods in free-living conditions, providing ecologically valid measures of gait quality that are strongly associated with future falls ^15^. Despite growing interest in sleep as a modifiable factor for healthy ageing, few studies have examined how self-reported sleep problems relate to objectively quantified daily-life gait quality and prospectively recorded fall incidence within the same cohort. The present study therefore investigated the relationship between self-reported sleep problems and fall risk in older people using complementary outcomes, by examining associations with daily-life gait characteristics and prospective fall incidence over one-year follow-up.

## METHODS

### Study design

This study is a secondary observational analysis of data from 758 people aged 65 and over who participatedin the StandingTall ^16^ and StandingTall+ ^17,18^ randomised control trials (RCTs). Although both trials evaluated exercise-based interventions, the current analyses focus on observational associations between sleep problems, gait quality, and fall incidence.

### Participants

Both studies had similar inclusion criteria in that participants had to be living in the community, independent in activities of daily living, able to walk household distances without a walking aid, have sufficient English language skills to understand the study procedures, be willing to provide informed consent and comply with the study protocol. Exclusion criteria were cognitive impairment defined as a Pfeiffer SPMSQ score <8 ^19^, current participation in a fall-prevention program, progressive neurological disease or any other unstable or acute medical condition precluding exercise. Participants in the StandingTall RCT were aged 70 years or over, while StandingTall+ RCT participants were aged 65 years or over and be at high fall risk, as defined by a history of falls in the past 6 months, a self-reported concern about falling, or aged 80 years or over. All participants provided informed consent, and ethics approval for both studies was obtained from the University of New South Wales Human Research Ethics Committee.

### Measures

Self-reported sleep was derived from the PHQ-9 sleep item (question 3) obtained at baseline: “*Over the last 2 weeks, how often have you been bothered by trouble falling asleep, staying asleep, or sleeping too much?*”, scored “*0, not at all*”, “*1, several days*”, “*2, more than half of the days*” or “*3, nearly every day*”. For primary analyses, this variable was collapsed into a binary indicator to indicate the presence of sleep problems: no sleep problems (score = 0) versus any sleep problems (score ≥ 1). This cut point had a sensitivity of 82.5% and a specificity of 84.5% for insomnia in a primary care setting ^20^.

Daily-life gait characteristics were assessed at baseline using a McRoberts activity monitor (McRoberts: The Hague, The Netherlands) worn continuously for one week during waking hours. Walking bouts were identified using validated algorithms and gait characteristics were derived to estimate a participant’s mean, or habitual, walking speed and gait quality composite score. The gait quality composite captures characteristics of gait variability, stability, and coordination during daily life and reflects dynamic balance control rather than overall walking capacity. This measure has previously been linked to falls and fall risk ^21^.

Falls and injuries from falls were prospectively recorded over 12 months using monthly digital fall calendars. An injurious fall was defined as a fall resulting in physical injury, including bruising, sprain, fracture, or injury requiring medical attention. Follow up phone calls were conducted when calendars were not returned.

Covariates included age in years, sex, body mass index (BMI), and years of education. Cognitive performance was assessed at baseline using the Montreal Cognitive Assessment (MoCA), and executive function was assessed using the Trail Making Test parts B minus A ^22^. A pain indicator was derived by collapsing self-reported pain locations (neck, shoulder, back, hip, knee, feet, other) into a binary variable (any pain vs none). Participants also self-reported the use of sleeping medications (yes/no).

### Statistical analysis

All analyses were conducted in RStudio. Associations between the binary sleep-problem exposure and baseline daily-life mobility outcomes were examined using linear regression. Fall incidence and injurious falls over 12 months were analysed using zero-inflated negative binomial regression models using *glmmTMB* with follow-up duration included as an offset. Zero-inflated models were selected based on evidence of overdispersion and excess zeros and provided the best fit compared with Poisson and standard negative binomial models.

Minimally adjusted models corrected for treatment allocation in the parent trial, while fully adjusted models further included a priori covariates selected to account for demographic and health-related confounding and to reflect established correlates of gait and falls: age, sex, BMI, pain, cognitive performance, executive function, and use of sleeping medications. To explore the contribution of gait quality to associations between sleep problems and falls, models were additionally adjusted for gait quality. Estimated marginal means and between-group differences (sleep problem vs no sleep problem) were derived from fitted models using *emmeans*, with 95% confidence intervals. Model assumptions were checked using residual diagnostics, Q-Q plots, and the *performance* package.

## RESULTS

Participants were on average 75.8 years old (standard deviation, SD = 5.9), and 69.3% were women (Table 1). Sleep problems were reported by 43.9% of participants (Figure 1), and 9.0% reported using sleep medication. Average habitual walking speed was 0.91 m/s (SD = 0.18), and the gait quality composite score was 0.48 (SD = 0.76). Over 12 months, 272 people experienced a fall (35.9%), 131 fell at least twice (17.3%), and 212 (28.0%) sustained an injury from a fall. The fall incidence rate was 0.92 falls per year (SD = 1.49), and the injurious fall incidence rate was 0.44 injurious falls per year (SD = 0.91).

**Table 1:**
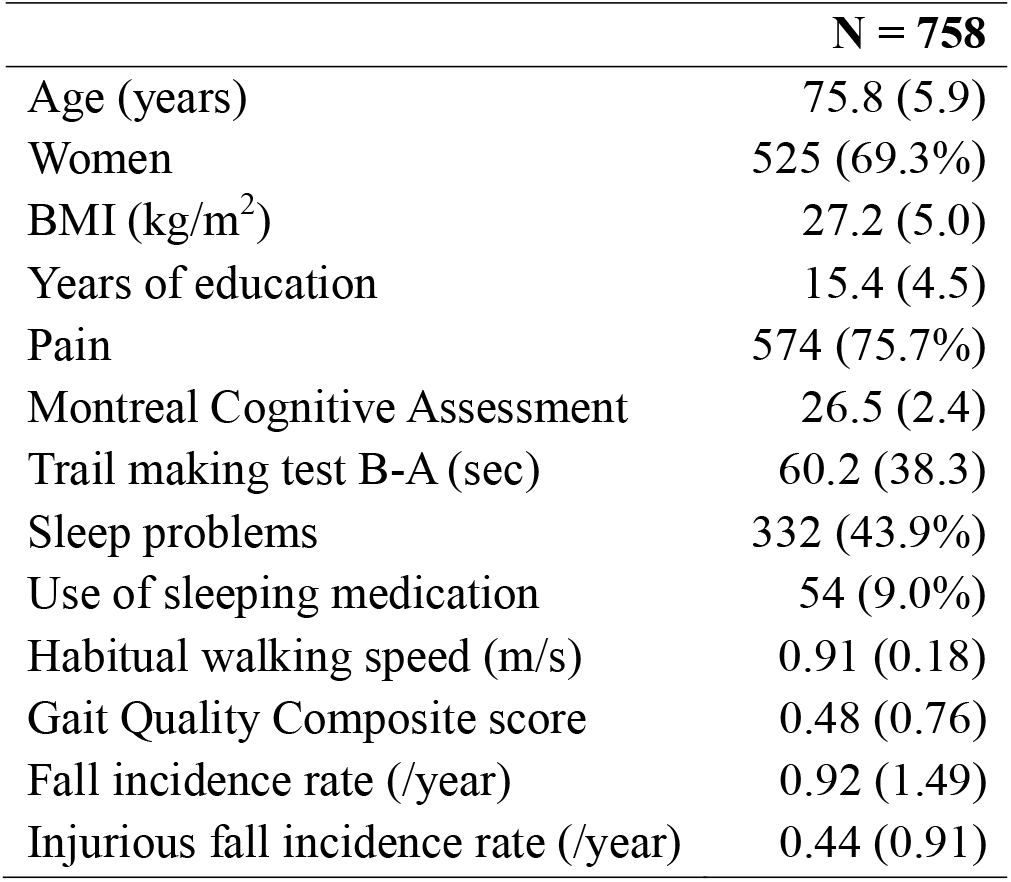
Demographic characteristics of the participants. Values are means (standard deviation) or N (%) unless stated otherwise

**Figure 1:**
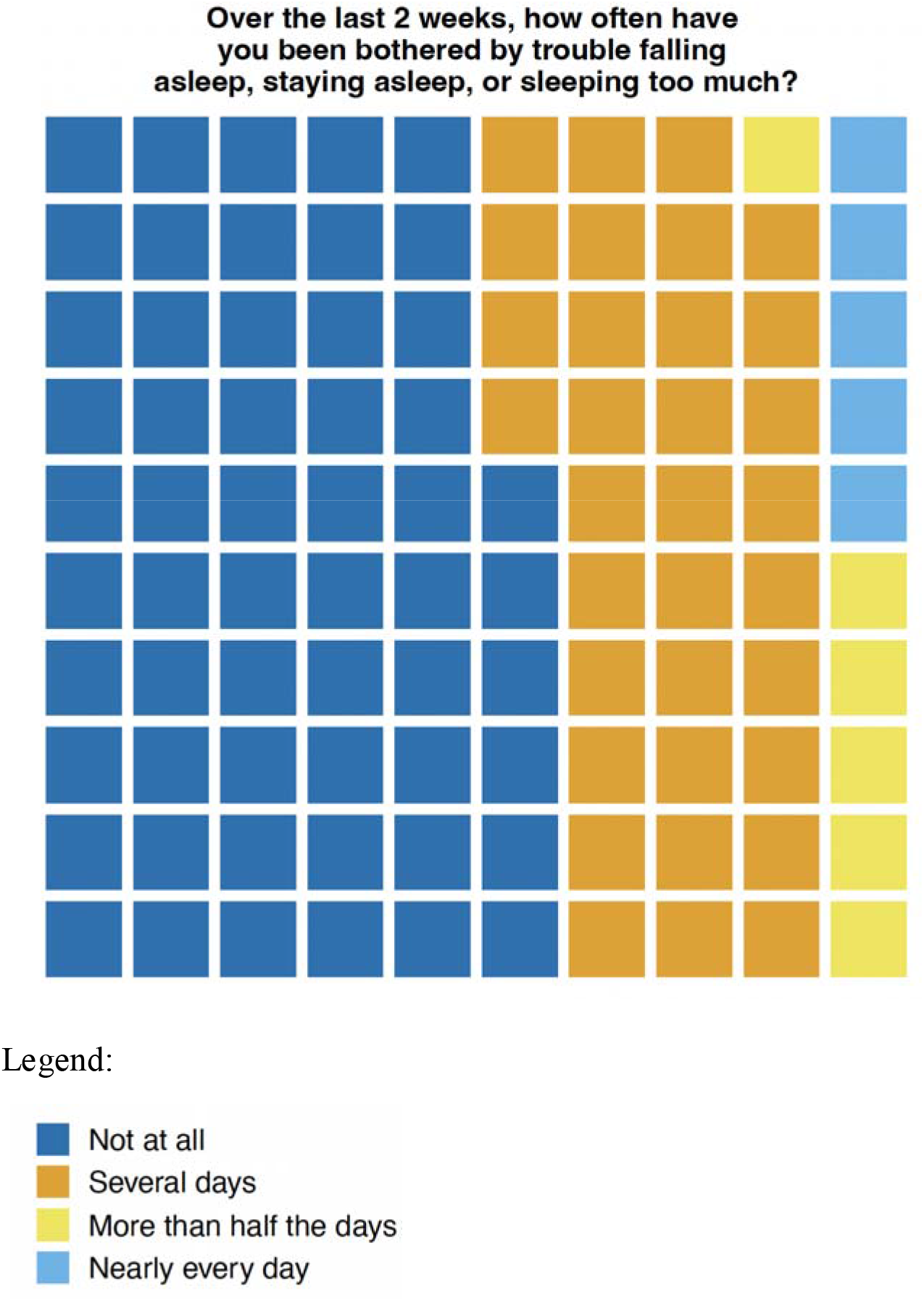
**Self-reported sleep** (% of participants, N = 758)

### Sleep and walking

People who reported sleep problems did not have a slower habitual walking speed compared to those who did not report sleep problems (Figure 2; minimally adjusted β = −0.0045 m/s, 95% confidence interval = −0.0301 to 0.0211; full adjusted β = 0.0017 m/s, 95% confidence interval = −0.0247 to 0.0280). However, they had a lower 0.15 point gait quality (unadjusted β = −0.15, 95% confidence interval = −0.26 to −0.04; adjusted β = −0.15, 95% confidence interval = −0.27 to −0.03), corresponding to a small effect size (Cohen’s d = −0.20).

**Figure 2:**
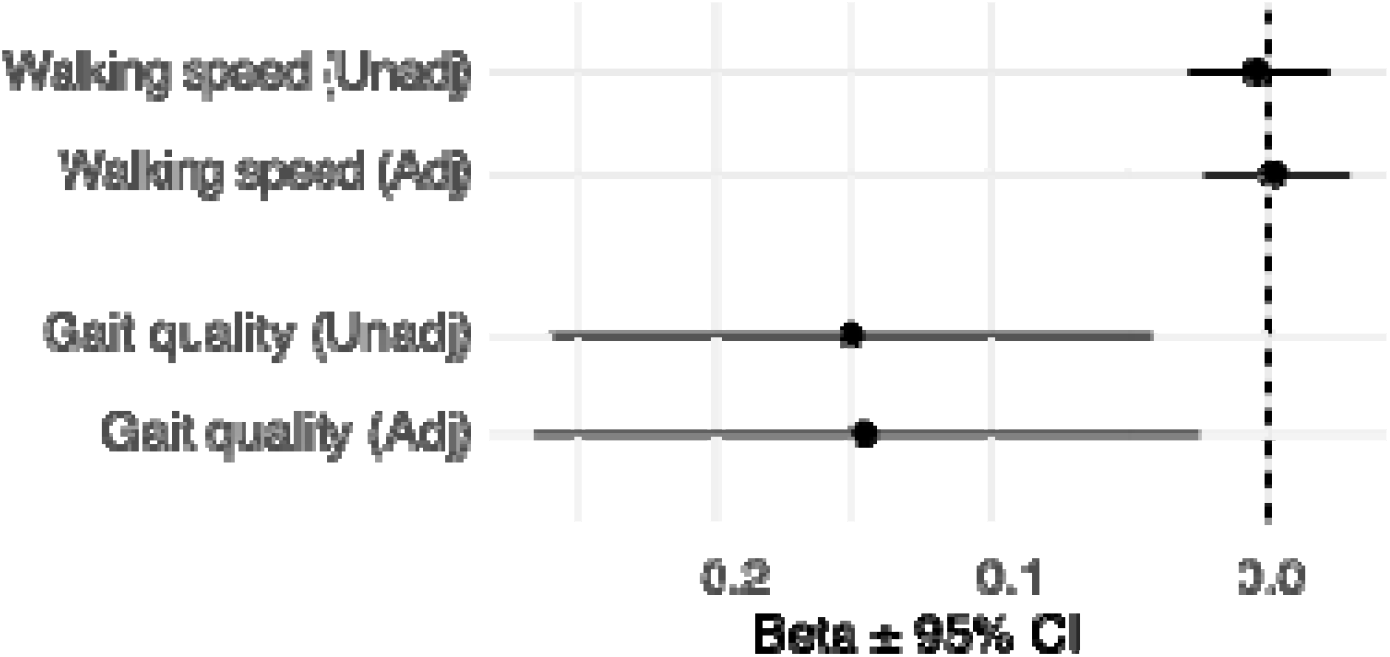
Association between sleep problems and gait.

### Sleep and falls

People who reported sleep problems had a higher incidence rate of falls and injurious falls (Figure 3). Specifically, sleep problems were associated with a 42-55% higher incidence rate of total falls (minimally adjusted incidence rate ratio (IRR) = 1.55, 95% confidence interval = 1.21 to 2.00; fully adjusted IRR = 1.42, 1.07 to 1.90) and a 45-50% higher incidence rate of injurious falls (minimally adjusted IRR = 1.45, 1.09 to 1.93; fully adjusted incidence rate ratio = 1.50, 1.07 to 2.10).

**Figure 3:**
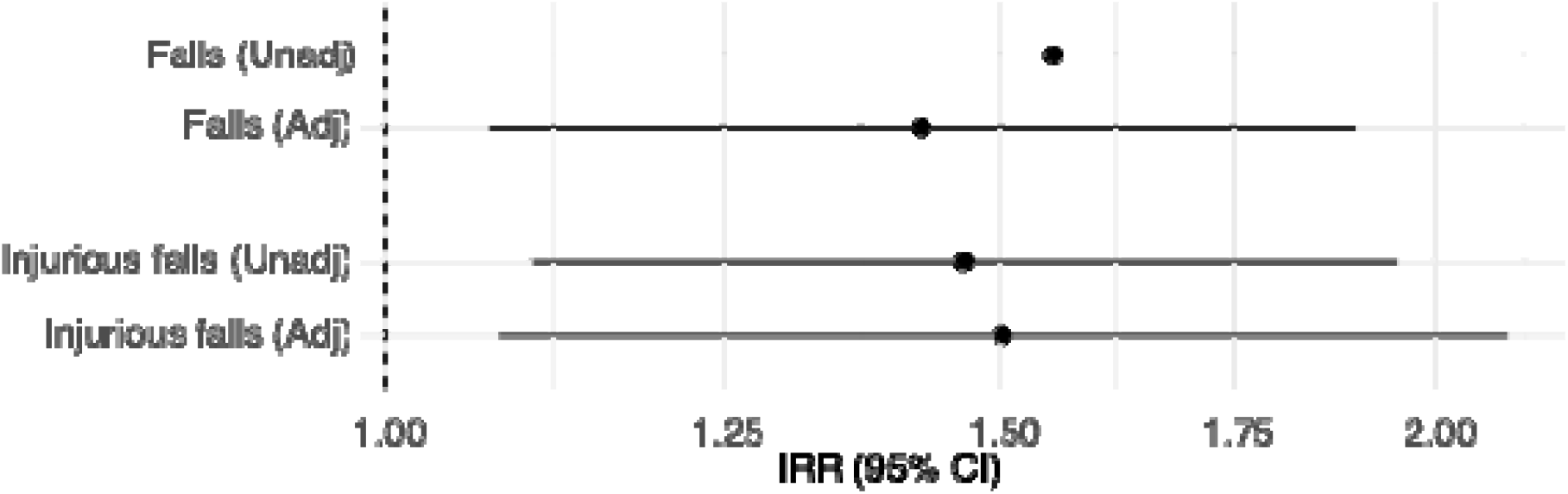
Association between sleep problems and falls.

Further adjustment for gait quality attenuated the associations between sleep problems and falls, with IRRs reduced to 1.34 (95% CI 1.02 to 1.76) for falls and to 1.44 (95% CI 1.03 to 2.02) for injurious falls, suggesting that a difference in gait quality accounts for 10 to 16% of the association between sleep problems and fall incidence.

## DISCUSSION

Sleep problems were common among older people living in the community, with 42% reporting poor sleep on several or more days. Sleep problems were not associated with differences in habitual daily-life walking speed. However, those reporting sleep problems had lower gait quality in daily life, corresponding to a small effect size comparable to differences typically observed between fallers and non-fallers, and to effects reported for medication or exercise interventions ^15^. Sleep problems were also associated with a 42 to 55% higher incidence rate of falls and fall-related injuries. These associations remained after adjustment for pain, cognition, and sleeping medication use, indicating that self-reported sleep problems identify a group of older people with elevated fall risk that is not fully captured by conventional risk factors. These findings highlight the need to consider sleep problems in fall risk management, as well as to consider fall risk in older people presenting with sleep complaints.

Sleep is known to affect functions relevant for safe mobility, including attention, information processing, and motor control ^6^. Such processes are critical for everyday walking, which requires continuous monitoring of the environment, divided attention, and rapid responses to perturbations ^9^. In older people, who often operate closer to their physiological limits, even subtle impairments in these systems may impair gait quality without necessarily impacting walking speed, as observed in the current study. This pattern is consistent with the distinction between gait quality as a marker of balance control during everyday walking, and habitual walking speed as a reflection of overall locomotor capacity and behavioural choice ^23,24^. The associations between sleep problems and both gait quality and falls persisted after adjustment for demographic variables, global cognition, executive function pain and the use of sleep medications, indicating that additional mechanisms may be involved. It is likely that sleep problems also influence fall risk through peripheral or neuromuscular processes. Poor sleep has been linked to reduced muscle function, impaired force generation, slower neuromuscular responses, and altered sensorimotor integration ^25-27^. These factors are directly relevant for maintaining balance during walking and for responding effectively to slips, trips, or other perturbations ^28^ and deserve further exploration.

Adjustment for gait quality only modestly attenuated the associations between sleep problems and both falls and injurious falls, with reductions of approximately 10-16%. This suggests that gait quality contributes to, but does not fully explain, the relationship between sleep problems and falls. Falls are complex events that arise from temporary states of vulnerability rather than sustained impairment ^29^, and sleep-related fall risk may also operate through transient states such as fatigue, daytime sleepiness, slowed reaction time, or impaired hazard perception. An alternative, and not mutually exclusive, explanation is that the relationship between sleep and mobility is dynamic rather than static. Sleep quality varies from night to night ^6^, and its effects on gait and balance may therefore fluctuate accordingly. Single baseline assessments may therefore underestimate the contribution of short-term changes in sleep to periods of heightened vulnerability to falls. Future studies that explicitly examine temporal relationships between sleep, daily-life gait characteristics, and balance control are needed to clarify these mechanisms.

Several limitations should be considered. First, sleep was assessed using a single self-reported item that captures perceived sleep problems but does not distinguish among specific sleep disorders, sleep duration, or objective sleep characteristics. While this measure reflects clinically relevant sleep complaints of insomnia, hypersomnia and sleep disruptions, future studies incorporating objective or multidimensional sleep assessments may provide greater mechanistic insight. Second, the observational design precludes causal inference. Although associations between sleep problems, gait quality, and fall incidence were robust and persisted after adjustment for multiple confounders, causality cannot be established, residual confounding cannot be excluded, and potential mediation pathways were not tested. Future studies are needed to determine whether improvements in sleep translate into measurable changes in gait stability and fall risk. Third, although daily-life gait quality provides ecologically valid information, it remains a composite marker and does not isolate which specific gait domains are most sensitive to sleep-related impairment.

From a clinical perspective, the findings suggest that sleep problems may be an important and under-recognised marker of fall risk among older people. Current *World Guidelines for Falls* focus on established risk factors, such as balance impairment, frailty and medication use ^4^.

Our findings indicate that routine enquiry about sleep health could complement these existing fall risk assessments, particularly in people who do not present with any other clear risk factors. Conversely, when older people present with sleep problems, consideration of fall risk may be warranted. While causality cannot be inferred from this observational study, addressing sleep problems using established non-pharmacological approaches, such as sleep hygiene education or cognitive behavioural strategies ^30^, and carefully reviewing sleep-related medication use ^31^, may offer a complementary avenue to support mobility and fall risk management.

In conclusion, self-reported sleep problems were associated with impaired daily-life gait quality and higher incidence of falls and injurious falls in community-living older people. These findings suggest that sleep problems may contribute to fall risk partly through alterations in balance control rather than reductions in walking speed. Integrating assessment of sleep problems with objective measures of real-world mobility may improve identification of older people at increased risk of falls and inform more comprehensive prevention strategies. Further studies combining objective sleep assessment with continuous monitoring of gait and balance over time will be essential to disentangle central, peripheral, and temporal mechanisms linking sleep to falls and to better understand how fluctuations in sleep contribute to periods of heightened vulnerability.

## Author contribution statement

KSvS conceived the idea for the analysis. KSvS, AV, RK, KS and KD designed the analyses. JB and KSvS prepared the dataset. KSvS performed the statistical analyses and drafted the manuscript. AV, RK, KS, JB and KD contributed to the interpretation of the results and critically revised the manuscript. All authors approved the final version.

## Acknowledgements

The authors thank the participants of the StandingTall and StandingTall+ RCTs for their time and commitment. We also acknowledge the research staff and students involved in data collection, participant support, and management of the wearable sensor data, as well as the broader StandingTall research team for their contributions to study delivery.

## Competing interest

None disclosed.

## Data availability

The datasets generated and/or analysed during the current study are not publicly available due to restrictions imposed by the ethics approval and participant consent, but are available from the corresponding author after an internal approval process.

## Notes

### Competing Interest Statement

The authors have declared no competing interest.

### Clinical Trial

ACTRN12615000138583 And ACTRN12619000540112

### Funding Statement

The original studies were funded by Australian National Health and Medical Research Council grant APP1084739 and APP1139673, Gandel Philaptropy, and NeuRA Foundation. The funders did not have a role in study design; in the collection, analysis, and interpretation of data; in the writing of the report; and in the decision to submit the paper for publication.

### Author Declarations

Human Research Ethics Committee of University of New South Wales gave ethical approval for this work (HC14266 and HC17977).

